# Unmasking the conversation on masks: Natural language processing for topical sentiment analysis of COVID-19 Twitter discourse

**DOI:** 10.1101/2020.08.28.20183863

**Authors:** Abraham C. Sanders, Rachael C. White, Lauren S. Severson, Rufeng Ma, Richard McQueen, Haniel C. Alcântara Paulo, Yucheng Zhang, John S. Erickson, Kristin P. Bennett

## Abstract

In this exploratory study, we scrutinize a database of over one million tweets collected from March to July 2020 to illustrate public attitudes towards mask usage during the COVID-19 pandemic. We employ natural language processing, clustering and sentiment analysis techniques to organize tweets relating to mask-wearing into high-level themes, then relay narratives for each theme using automatic text summarization. In recent months, a body of literature has highlighted the robustness of trends in online activity as proxies for the sociological impact of COVID-19. We find that topic clustering based on mask-related Twitter data offers revealing insights into societal perceptions of COVID-19 and techniques for its prevention. We observe that the volume and polarity of mask-related tweets has greatly increased. Importantly, the analysis pipeline presented may be leveraged by the health community for qualitative assessment of public response to health intervention techniques in real time.

## 1 Introduction

Social media provides a rich corpus of text characterizing in real time the daily happenings and current events within our communities. As such, it has potential utility for individuals and entities wishing to keep their fingers on the pulse of both social and public health issues. Mask-wearing during the COVID-19 pandemic falls into both categories, as the consensus in the scientific community that wearing masks is key to controlling the spread of the SARS-CoV-2 virus^1^ has been met with non-negligible resistance for various sociopolitical reasons. Research avenues investigating this mask usage discrepancy are increasingly relevant in light of both the evolution of the coronavirus pandemic into a border-independent global crisis and the extent to which public perceptions of the virus have changed over time.

### Background and Related Works

In the pandemic-era reality that has evolved in 2020, social distancing has become the necessary norm, and it is known that social media is playing a bigger role than ever in keeping people connected and informed.^2^ Several mainstream social media platforms have seen usage spikes amongst English-speakers since the onset of the pandemic.^3^ In keeping with the stimulation of social media activity observed to accompany disease outbreak events, a body of literature has emerged over the past decade that looks specifically at how trends in online activity and discourse can help inform epidemiological models.^4^ In conjunction, a suite of programming frameworks and models drawing on data harvested from Twitter have been developed to answer specific research questions about viral trends and their societal impacts.^5–7^

### Major Contributions

This analysis aims to provide insight into the broadscale conversation surrounding mask-wearing evolving on Twitter between March and July of 2020, when infection rates initially spiked in the United States, Europe, and other regions throughout the world. To this end, we develop a novel pipeline employing state-of-the-art natural language processing (NLP) techniques in order to systematically characterize Twitter discourse about and public attitudes towards the topic of mask usage during the COVID-19 pandemic. Specifically, we collect and analyze a comprehensive sample of coronavirus-related tweets textually related to mask-wearing. We employ clustering techniques to organize these tweets into fifteen high-level themes and fifteen specific topics within each theme, then perform sentiment analysis on the entire corpus, and also on each theme and topic, across a five-month period. We then apply an abstractive text summarization model using NLP to automatically interpret and describe the subject of the conversation occurring within each theme and topic cluster. We use data visualization and statistical analyses to examine trends in sentiments and divisiveness of the clusters.

Our pipeline is distinct from others recently developed for COVID-19-related information characterization. While other works have primarily drawn from unfiltered Twitter corpora, or alternatively, from manually-annotated datasets specific to a particular hypothesis, we chose to compromise between the two approaches by refining an index of tweets strictly related to both COVID-19 and masks based on text-based keyword identification. With this semi-selective approach, we highlight the thematic trends that manifest organically in the tweets we have collected, while also ensuring that the global English-speaking conversation surrounding mask-usage during the pandemic is represented.

We find two central, co-occurring trends in the English-speaking Twitterverse by means of the presented pipeline. First, Twitter discourse surrounding mask-wearing within our curated dataset is concluded to grow consistently polarized over time, irrespective of the high-level topic into which it is clustered. Moreover, we find evidence to suggest that sentimentality related to masks and mask-use as expressed on Twitter grew increasingly negative as the pandemic progressed. Cumulatively, we concur that a qualitative, semantic Twitter-based analysis pipeline is capable of revealing striking insights into public responses to the pandemic. We hope that the methods developed here can evolve into tools to help provide rapid real-time assessment of public health measures to inform future interventions.

## 2 Methods

### 2.1 Data Collection

We used the Twitter streaming API^8^ to collect 189,958,459 original tweets filtered by keywords loosely associated with COVID-19 ^1^ over a five month period beginning on March 17^th^, 2020 and ending on July 27^th^, 2020. We restricted our filter to English-language tweets via the streaming API language filter parameter, and discarded any retweets during this time period. Twitter’s API provides access to a representative random sample of approximately 1% of all tweets in near real time, and it has been shown that samples obtained via the API reflect the general content generation patterns of the Twittersphere accurately.^9^ We stored all collected tweets in Elasticsearch^10^ indices for efficient search and retrieval. Using Elasticsearch, we further filtered our corpus of collected tweets by the criteria that a tweet must include at least one keyword indicating it is strongly associated with COVID-19 and at least one keyword indicating it is strongly associated with mask-wearing. This filter yielded a corpus of 1,013,039 tweets for our analysis.

Although we analyze temporal elements which could be interpreted in a regional context, we chose not to filter our dataset geographically. Users who share their device location represent less than 1% of the Twitter population.^11^ Additionally, studies have reported 34% of user-specified profile locations are unusable,^12^ and the remainder may not reliably represent tweet origin.^13^ We made this decision to avoid any biases that may be present in geotagging Twitter usership and to preclude the risk of inaccuracies inferring tweet location from profile information.

The collected corpus of tweets and the full source code for the data collection analysis pipeline are publicly available at https://github.com/TheRensselaerIDEA/COVID-masks-nlp. In compliance with the Twitter content redistribution policy^2^, we only provide the tweet IDs corresponding to the collected tweet text used in this work.

### 2.2 Analysis Pipeline

We develop an analysis pipeline to extract, label, summarize, and present the themes, topics and sentiment present in our tweet corpus using state-of-the-art natural language processing tools. While we use it here for analysis of our corpus pertaining to mask-wearing, our methods can be applied to any dataset of text documents. We have included an online supplement ^3^ containing additional details on implementation decisions and software packages used.

#### Step 1: Retrieval & Sampling

The first step in the analysis pipeline is the retrieval of a representative random sample of tweets from the corpus. We chose *N* = 100, 000 as our sample size for this study, and restricted sampled tweets to those created within the range of March 1^st^, 2020 to August 1^st^, 2020 - a sample space of 1,012,815 tweets. Once retrieved, all tweets are cleaned by removing URLs and non-punctuation characters and then normalizing all whitespace character sequences to single spaces.

#### Step 2: Embedding & Sentiment Scoring

After retrieving and cleaning the sample, each tweet is embedded into a 512-dimensional vector space using the transformer^14^ implementation of Google’s Universal Sentence Encoder.^15^ The vector that represents each tweet is given by the sum of the contextual word representations at each position of the transformer encoder output. Semantically similar tweets are grouped together in the resulting embedding space, where cosine similarity provides a metric of how close two tweets are in meaning.

To assess tweet sentiment, each tweet is also scored using the VADER algorithm - a social-media-centric, lexicon-based sentiment characterization approach.^16^ VADER provides a compound polarity score between −1 (most negative) and 1 (most positive). For graphical representations, we use the authors’ recommended threshold of *±*0.05 to discretize the score where *s ≤ −*0.05 is negative, *−*0.05 *< s <* 0.05 is neutral, and *s ≥* 0.05 is positive.

#### Step 3: Clustering & Subclustering

Next, we apply k-means in the embedding space to create a two-level cluster hierarchy - the corpus is grouped into *k* primary clusters and each primary cluster is then grouped into *k*_*c*_ subclusters. We interpret the primary clusters as representing high-level discussion themes and the subclusters as specific topics within each theme. We re-order the cluster numbers 1 through *k* and subcluster numbers 1 through *k*_*c*_ by average sentiment score, with 1 being the most negative. To select the optimal number of primary clusters and subclusters, we performed a computational study of the k-means objective function across a range of choices for *k* and *k*_*c*_. As documented in our supplement, we selected *k* = 15 and *k*_*c*_ = 15 since these values provided a good balance between cluster quality and avoidance of topical redundancy.

We then use t-Distributed Stochastic Neighbor Embedding (t-SNE)^17^ to project the clustered embedding space into two dimensions for presentation. In Figure 1, the cluster and subcluster scatterplots use coordinates in R2 given by t-SNE. The primary cluster plot is color-coded by cluster assignment and the subcluster plots are color-coded by subcluster assignment. The black points represent the cluster and subcluster centers.

**Figure 1:**
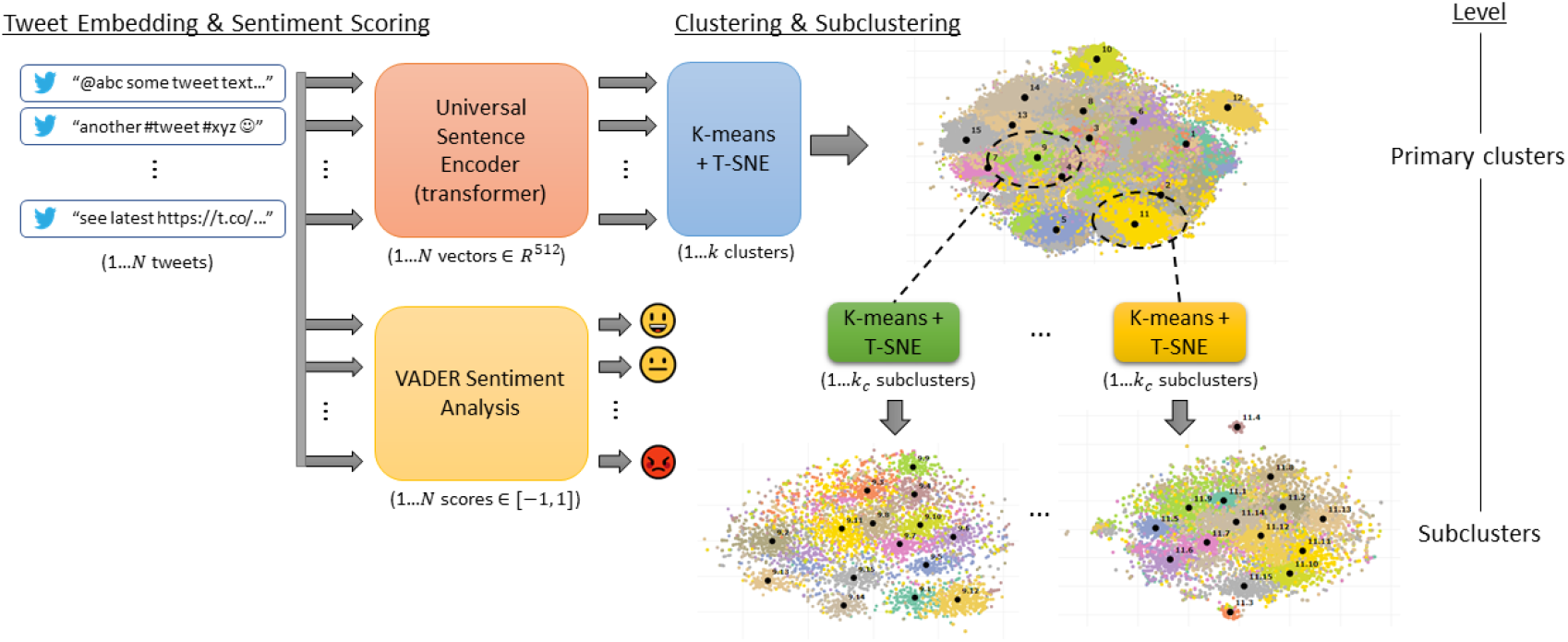
K-means is used to cluster the tweets in their embedding space. A two-level cluster hierarchy is created by applying k-means again to each cluster.

**Figure 2:**
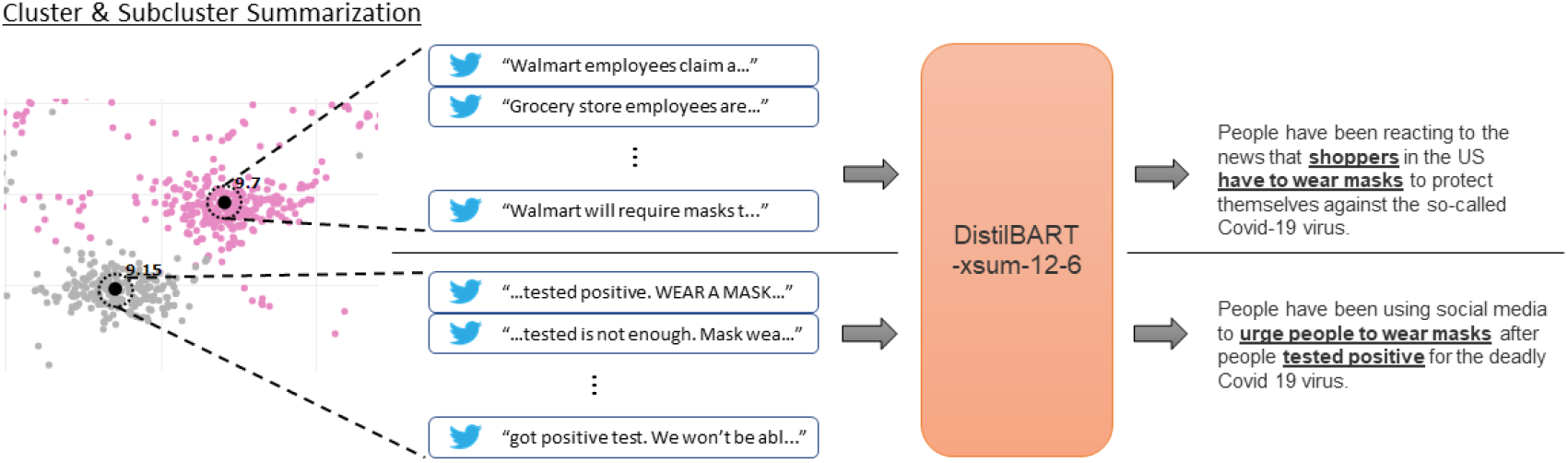
The tweets embedded nearest the subcluster center (shown as a black dot) are used to create the input “article” for DistilBART to summarize.

We recognize there are many approaches for topic extraction from short text. Our choice to embed tweets with Universal Sentence Encoder and cluster them with k-means in the embedding space was intended to capture context-sensitive representations of tweet text. While our method proves effective, a range of other methods may be used to similar ends. Wang et al. report that k-means combined with sentence embeddings performs comparably to traditional methods such as LDA^18^ and TF-IDF on unsupervised Twitter topic modeling, with XLNet^19^ and Universal Sentence Encoder outperforming other models on a variety of metrics.^20^

#### Step 4: Cluster & Subcluster Labeling

We find keywords that both describe and differentiate the discussion within each cluster and subcluster, and use these keywords as labels. To do this, we compute relative frequencies for words across each cluster, ignoring stopwords and non-alphanumeric characters. Using the relative frequencies, we score each word according to its contribution to the Kullback-Leibler divergence between the word distribution of the cluster and the word distribution of the entire corpus sample: 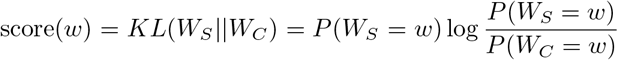. Here, *W*_*C*_ and *W*_*S*_ are the word probability distributions for the corpus sample and sub-sample (cluster), respectively. Subclusters are labeled in the same manner, with the parent cluster taking the place of the corpus sample. Additional illustration of the labeling method is included in the supplement.

A single label representing the corpus sample is computed using the eight words with the highest overall frequencies. For each cluster and subcluster, we select the three words with the highest scores and concatenate them to create theme and topic labels respectively. To avoid reuse of keywords across labels, cluster labels cannot contain keywords that exist in the corpus sample label, and subcluster labels can not contain keywords that exist in the parent cluster label.

#### Step 5: Cluster & Subcluster Summarization

To augment human interpretations of each cluster and subcluster, we generate summaries using DistilBART, an abstractive summarization model from the HuggingFace Transformers^21^ package based on Facebook’s BART^22^ model. While the labels provide a quick description of the type of discussion happening within a cluster or subcluster, the one-to-three sentence summary produced by this process conveys this information in a much more meaningful way. We use a DistilBART instance fine-tuned on the extreme summarization (xsum) task^23^ which aims to generate concise summaries of articles without relying on extractive summarization strategies. For each subcluster, we generate the input “article” for DistilBART to summarize by concatenating the text of 20 tweets which are embedded nearest to the subcluster center. For each cluster, we generate the input by concatenating all of the model-generated summaries of its subclusters. We performed a qualitative study to assess two additional strategies for summarizing the clusters, and selected this approach as we found it least prone to misrepresentation of cluster themes. More detail on this study have been included in the supplement.

It should be noted that the xsum dataset used to fine-tune DistilBART is comprised of news articles from the British Broadcasting Corporation (BBC) and their corresponding summaries. Multi-tweet summarization is an admittedly different task domain with noisier, less consistent text. We consider our application of DistilBART in this domain to be proof-of-concept, with the logical next step of fine-tuning on a human-annotated xsum-like dataset for Twitter in keeping with the original methods of Narayan et al.^23^

### 2.3 Sentiment Analysis

#### Divisiveness in Sentiment

In order to better understand the sentiment profile of the tweet clusters, we developed a divisiveness score to assess the present level of polarization in tweet sentiment. The score is given by a real number such that polarized samples with little neutral sentiment are given a positive score, while samples with consensus (unimodally concentrated on a single sentiment category) are given a negative score. Otherwise, in the case where sentiment is uniformly distributed across categories, samples have a score of zero. The score is based on the Sarle’s Bimodality Coefficient^24^ (*BC*) with an added correction through a weighted average with the *BC* of the uniform distribution, and then a logit transformation. This weighting counterbalances the large variance of the *BC*, based on the skewness and kurtosis for small samples,^25^ so that such samples with little information are considered to still have uniformly polarized sentiment.

## 3 Results

We examine the tweet volume and sentiment concerning masks for the entire sample. Table 2 contains sentiment average, overall divisiveness, and trends in divisiveness for each cluster for tweets from March to July 2020. Cluster interpretations in Section 4 further illustrate the nature of the mask discourse.

**Table 1:** Filter criteria we used to identify tweets that are related to both COVID-19 and mask-wearing. A tweet must contain at least one keyphrase in both categories to be included.

| Keyphrases related to COVID-19 | Keyphrases related to mask-wearing |
| --- | --- |
| “ncov”, “sars-cov-2”, “covid”, “covd”, “covid19”, “corona”, “virus”, “coronavirus”, “koronavirus”, “wuhancoronavirus”, “kungflu”, “epidemic”, “pandemic”, “quarantine”, “lockdown”, “flatten the curve”, “flattenthecurve”, “cdc” | “mask”, “wearamask”, “masking”, “N95”, “face cover”, “face covering”, “face covered”, “mouth cover”, “mouth covering”, “mouth covered”, “nose cover”, “nose covering”, “nose covered”, “cover your face”, “coveryourface” |

**Table 2:**
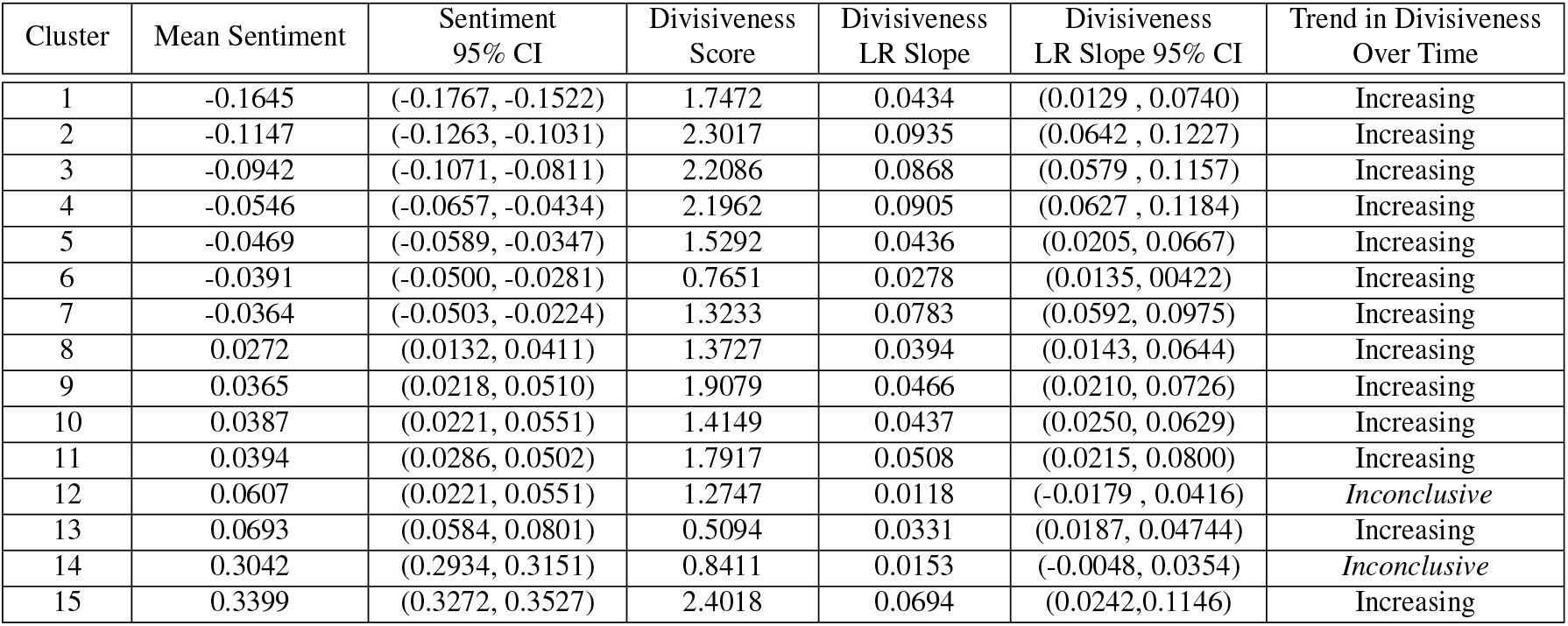
Average sentiment scores, divisiveness scores, and regression line slopes with 95% confidence intervals, and qualitative descriptions of time series trends. Clusters are listed in order of increasing sentiment score.

| Cluster | Mean Sentiment | Sentiment<br>95% CI | Divisiveness<br>Score | Divisiveness<br>LR Slope | Divisiveness<br>LR Slope 95% CI | Trend in Divisiveness<br>Over Time |
| --- | --- | --- | --- | --- | --- | --- |
| 1 | -0.1645 | (-0.1767, -0.1522) | 1.7472 | 0.0434 | (0.0129, 0.0740) | Increasing |
| 2 | -0.1147 | (-0.1263, -0.1031) | 2.3017 | 0.0935 | (0.0642, 0.1227) | Increasing |
| 3 | -0.0942 | (-0.1071, -0.0811) | 2.2086 | 0.0868 | (0.0579, 0.1157) | Increasing |
| 4 | -0.0546 | (-0.0657, -0.0434) | 2.1962 | 0.0905 | (0.0627, 0.1184) | Increasing |
| 5 | -0.0469 | (-0.0589, -0.0347) | 1.5292 | 0.0436 | (0.0205, 0.0667) | Increasing |
| 6 | -0.0391 | (-0.0500, -0.0281) | 0.7651 | 0.0278 | (0.0135, 0.0422) | Increasing |
| 7 | -0.0364 | (-0.0503, -0.0224) | 1.3233 | 0.0783 | (0.0592, 0.0975) | Increasing |
| 8 | 0.0272 | (0.0132, 0.0411) | 1.3727 | 0.0394 | (0.0143, 0.0644) | Increasing |
| 9 | 0.0365 | (0.0218, 0.0510) | 1.9079 | 0.0466 | (0.0210, 0.0726) | Increasing |
| 10 | 0.0387 | (0.0221, 0.0551) | 1.4149 | 0.0437 | (0.0250, 0.0629) | Increasing |
| 11 | 0.0394 | (0.0286, 0.0502) | 1.7917 | 0.0508 | (0.0215, 0.0800) | Increasing |
| 12 | 0.0607 | (0.0221, 0.0551) | 1.2747 | 0.0118 | (-0.0179, 0.0416) | Inconclusive |
| 13 | 0.0693 | (0.0584, 0.0801) | 0.5094 | 0.0331 | (0.0187, 0.04744) | Increasing |
| 14 | 0.3042 | (0.2934, 0.3151) | 0.8411 | 0.0153 | (-0.0048, 0.0354) | Inconclusive |
| 15 | 0.3399 | (0.3272, 0.3527) | 2.4018 | 0.0694 | (0.0242, 0.1146) | Increasing |

Figure 3a shows the number of negative (red), neutral (yellow) and positive tweets (blue) per week. Clearly the volume and polarity of the discussion have dramatically increased starting in mid-June. Figure 3b shows the labels provided by the keyword analysis for each cluster, ordered from most negative sentiment to most positive sentiment. Figure 3c shows the weekly counts for tweets by sentiment for each week. Clusters 1-3 are the most negative clusters, which, as later detailed in Section 4, respectively discuss the topics of Donald Trump, individuals not wearing masks, and government mask and social distancing mandates.

**Figure 3:**
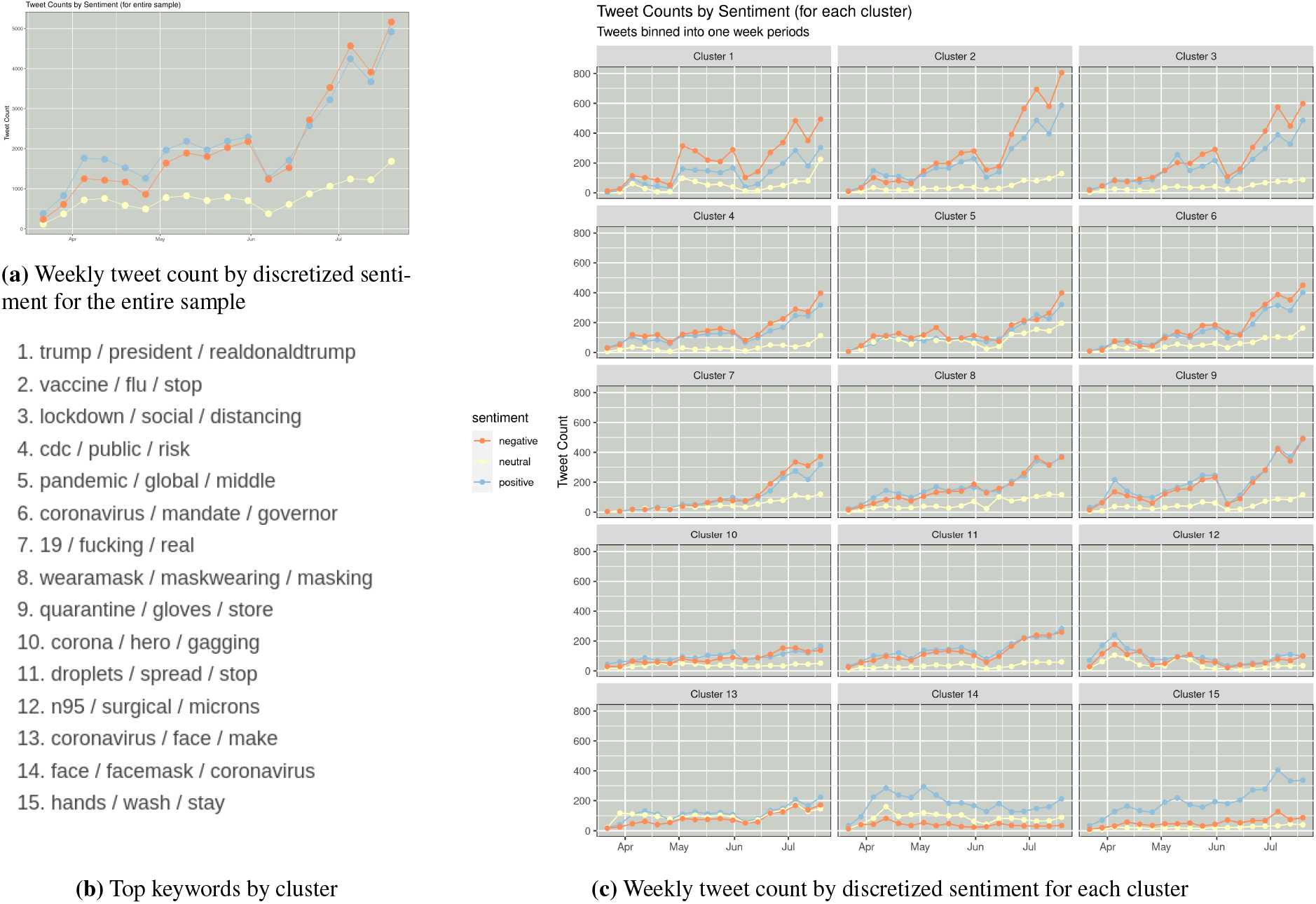
Sentiment over time, for the entire tweet corpus and for each cluster

### Cluster Divisiveness

To characterize the polarization of each topic cluster and the changes in polarization over time, we perform global and per-week analyses of the divisiveness scores for all clusters. For each cluster we compute divisiveness for each week, then run a linear regression of divisiveness against time; the results are shown in Table 2.

We see that for all clusters, except for Clusters 12 and 14, the confidence intervals for the slope of the fitted lines are entirely positive, indicating an increasing trend in divisiveness over time. However, no clusters display particularly steep trends, with the most significant one being Cluster 13 with a slope equivalent to only 0.0649% of the overall divisiveness score. All clusters are shown to be divisive; however, Clusters 6 and 13 possess the lowest divisiveness scores, while Clusters 2, 3 and 15 are shown to be the most divisive. Cluster 15 in particular is found to have the greatest divisiveness score. However, this result likely comes from a known fault of Sarle’s BC when handling heavily skewed distributions.^24^ In this case, the divisiveness score is likely incorrectly inflated due to the cluster distribution being heavily skewed towards positive sentiment, shown in Figure 3c. Clusters 2 and 3 then evidently come out to be the most polarizing out of all clusters presented, both also having comparatively large values for the fitted regression line slope with 95% certainty of increasing sentiment divisiveness.

### Variance in Sentiment Over Time

A one-way ANOVA was conducted for differences in mask-related sentiment across five consecutive months of early 2020 (March through July) using the complete tweet corpus, *N* = 1, 013, 039. The ANOVA was performed on the basis of observed normality of residuals, and with the caveat that a Breusch-Pagan test pointed to heterogeneity of variance between months. The test result indicated with significance (*p <* 10^*−*16^) the presence of at least one distinct difference in sentiment among the five pandemic months analyzed. A subsequent Bonferroni-corrected pairwise t-test further confirmed statistically significant differences in mean sentiment score across all months studied (*p <* 0.001). In light of this finding, we followed with Dunnett’s contrasts^26^ to compare the mean sentiment for each group to that of March, the earliest month in our dataset. The results concurred, at *α* = .05, that the mean sentiment scores computed for the months of April through July all differed significantly from that of March, at *p* = 0.0143 for April and *p* < 0.001 for all other months. We re-ran the Dunnett method with the alternative hypothesis that the mean sentiment for each month was greater than or equal to that of March. We failed to reject the null hypothesis of a decrease for each pairwise test. Taken together with the graphical trends found, we interpret this result to suggest an overall decrease in mean sentiment score related to masks and mask-use as of July 2020.

## 4 Cluster Interpretations

In this section, we select five clusters found to be particularly striking in content. We order the clusters by increasing overall sentiment score, report on the trends in sentiment and divisiveness metrics, and include the automatically-generated summary for each. We then provide manual annotations of the prominent themes that arise, by inspecting small samples of tweets lying near each of the fifteen subcluster centers within each cluster. We see that support for mask wearing and cluster sentiment do not necessarily correspond.

### Cluster 1: trump / president / realdonaldtrump (Overall Sentiment : −0.1645 ; Divisiveness : 1.7472)

#### DistilBart summary

*People have been reacting to news that President Donald Trump has refused to wear a face mask in public to protect himself from the deadly coronavirus pandemic*.

#### Interpretation

This cluster (shown in Figure 4) features Twitter users expressing a spectrum of attitudes towards U.S. President Donald Trump. Opinions specifically revolve around Trump’s handling of the COVID-19 pandemic in the United States. Distinctly, there exists an evident theme of frustration arising from observations that Trump has refused to wear a mask in public appearances, despite statements from public health officials encouraging the action. In complement, a sizeable positive discussion thread also exists concerning President Trump. A major theme observed here among the pro-Trump tweets is the impression that the media is biased against the president, and that this in turn fosters a public motive to exaggerate the virus.

**Figure 4:**
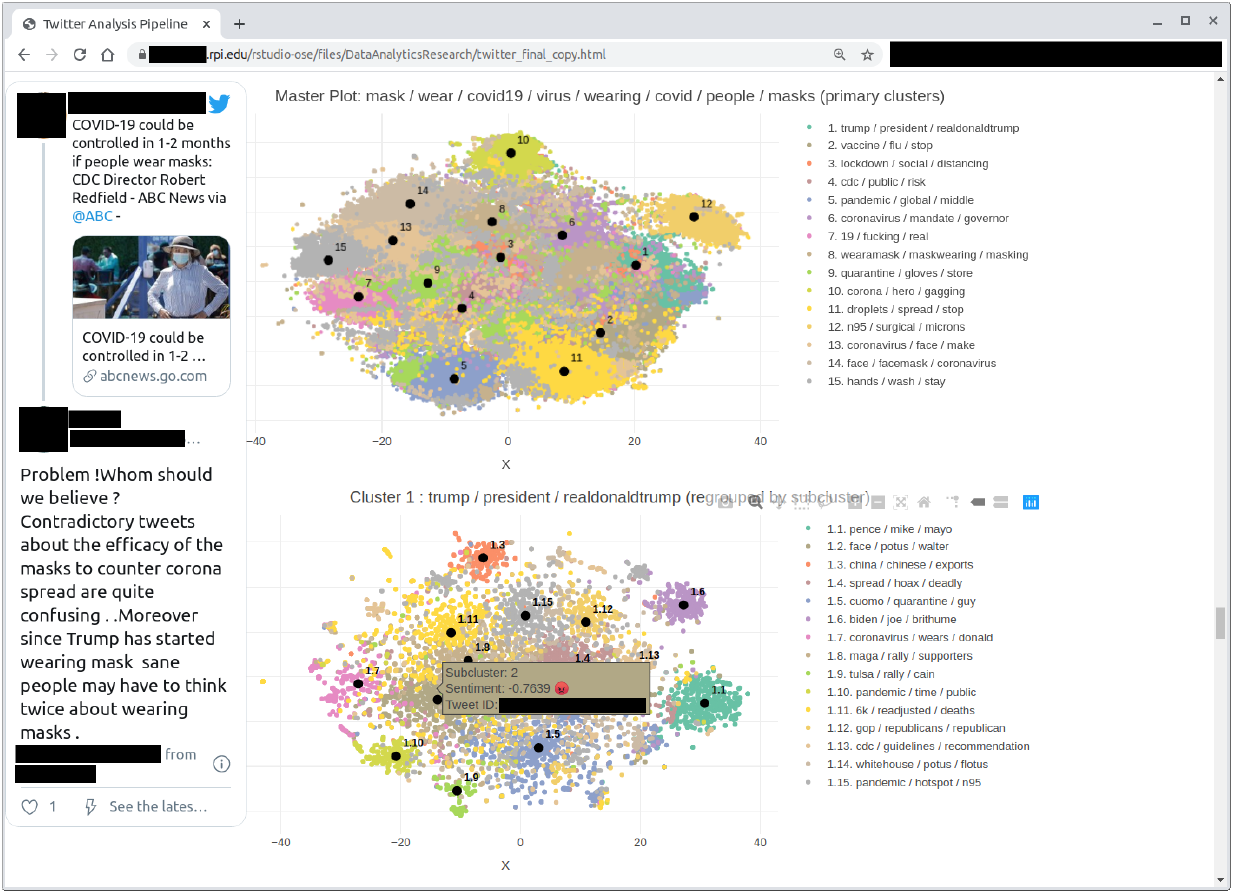
We have made available an interactive document containing the full listing of all clusters, subclusters, and automatically-generated summaries. Our interactive notebook containing results for all clusters can be found at https://therensselaeridea.github.io/COVID-masks-nlp/analysis/twitter.html

### Cluster 2: vaccine / flu / stop (Overall Sentiment : −0.1147 ; Divisiveness : 2.3017)

#### DistilBART summary

*Following the news that people in the US are being urged to wear face-covering masks to prevent the spread of a new virus that has killed more than 4,000 people in China*.

#### Interpretation

Cluster 2, “vaccine / flu / stop”, is a grim cluster in terms of its overall sentiment, and is distinctly polemical in its semantics. It is found that the majority of tweets sampled from this cluster complain about individuals who don’t wear masks. The dominant attitude towards masks is positive, despite the overall negative sentimentality computed for the cluster as a whole. In contrast with the more semantically upbeat “face / hands / stay” cluster (Cluster 15), the theme of death and dying is prevalent. The social nature of disease is a major motif (i.e. “Your actions affect all of us.”)

### Cluster 3: lockdown / social / distancing (Overall Sentiment : −0.0942 ; Divisiveness 2.3017)

#### DistilBART summary

*Following the news that the US government has ordered people to wear face masks in public to prevent the spread of the deadly Covid-19 coronavirus, people across the world have been reacting to the news on social media*.

#### Interpretation

Cluster 3 gives an indication of the societal turbulence arising from mask mandates, social distancing enforcement, and similar lockdown-related measures globally. Paradoxically, the overall average sentiment of −0.0941 computed for this cluster is borderline neutral. A strong racial emphasis is evident, with discourse focusing around protests of the #BlackLivesMatter movement, an international phenomenon co-occurring with the coronavirus pandemic mid-year. Several regions of Cluster 3 contain conversations about international responses to the virus, notably around the idea that mask-wearing to prevent the spread of disease agents is a long-standing cultural norm in some regions. In keeping with the slightly negative overall sentiment for this cluster, many of the tweets express sarcasm.

### Cluster 12: n95 / surgical / microns (Overall Sentiment : 0.0693 ; Divisiveness : 1.2747)

#### DistilBART summary

*News that a shortage of N95 respirator masks in the US is causing a worldwide shortage has been shared on social media*.

#### Interpretation

Discourse within Cluster 12 focuses on information about N95 masks and related forms of personal protective equipment (P.P.E.). The evolution of the conversation around the accessibility of medical resources over the timeline of the pandemic is clearly represented. One notable stream of discussion points to the presence of a debate over how effective cloth masks are as guards against infectious agents in comparison to surgical masks. The shortage of respirators experienced by the medical community in the United States is also referenced, as is a change in perspective on N95 masks from the U.S. CDC in the early days of the pandemic.

### Cluster 15: hand / wash / stay (Overall Sentiment : 0.3399 ; Divisiveness : 2.4018)

#### DistilBART summary

*Social media users have been sharing their tips and advice on how to prevent the spread of the deadly coronavirus*.

#### Interpretation

Our most positive cluster, “hand / wash / stay” is composed of tweets sharing tips on prevention measures for stopping the spread of COVID-19. There appears to be highly positive sentiment expressed towards masks and other PPE, and well-meaning admonitions such as “Wash your hands and socially distance!” are frequent. In contrast to other clusters, the Cluster 15 tweets contain little in the way of aggressive, sarcastic or antagonistic semantics. As such, to an extent, this cluster echoes the official messaging of the CDC and similar organizations.

## 5 Discussion

The objective of our analysis framework was to study the distribution of global mask-related social media discourse, the specific topics within this distribution, their sentimentality trends and how the latter have changed over time. In comparison to the official sources of COVID-19 infection and death rate data, the accessibility and sheer quantity of organic discourse played out over Twitter make this platform an invaluable source of information on public perception of mask usage during the coronavirus pandemic. The cumulative results of our pipeline point to the existence of two central, co-occurring trends in the English-speaking Twitterverse: consistently polarized Twitter discourse surrounding mask-wearing, and an accompanying overall increase in negative sentimentality.

While mask-wearing is a health-related issue, the politicization of mask-wearing is exposed in this investigation. The fact that the U.S. president holds such bearing in the global Twitter conversation about mask-wearing speaks to the degree to which sociopolitical dynamics hold sway over the public perception of the pandemic. The topic-sensitivity of the clustering approach we develop also opens doors for new health-related insights regarding COVID-19’s impact.

While our pipeline is effective, opportunities for improvement exist. Any component of the pipeline can potentially be replaced. For example, alternative clustering methods could be used. An open question arising from this research is how well VADER-computed sentiment estimations reflect public opinion in a semantic sense. In this work, we leverage lexicon-based sentiment analysis as a proxy for human attitudes and emotions, but we plan to incorporate a more holistic sentiment representation moving forward (e.g. one capable of detecting expressions of sarcasm).

Two important limitations of our summarization method should be noted. First, the BART-based decoder is a generative language model which creates summaries autoregressively by repeatedly sampling from next-word probability distributions over an entire vocabulary. For this reason, the output summaries are prone to factual inaccuracy in a manner which extractive summarization approaches are not. Second, large or irregularly shaped subclusters may be poorly represented by the tweets immediately surrounding the subcluster center. In these situations the generated summary may not be applicable to the entire subcluster. We accept these as limitations of the system when used with unannotated data, as is the case in our study. As such, we advise users of the pipeline to regard the summaries as context clues, and then use the notebook provided for further investigation.

## 6 Conclusion

In light of both the escalation of the pandemic into a global crisis and the extent to which the implications of the virus have changed in the public eye over time, semantic analyses such as we present are increasingly relevant as sources of information to the medical research community for a host of health-related considerations. As we see, mining Twitter data allows for the rapid summarization of opinions about empirically-supported disease prevention measures. Overall, we find that thematic clustering and visualization based on mask-related Twitter data can offer distinct insights into societal perceptions of COVID-19, complementary to findings from more traditional epidemiological data sources. With the aid of abstractive visualizations like the clustering techniques presented, acute estimations of what individuals are actually saying and feeling amidst the viral destruction can be made. As future work, we hope to evolve this pipeline into a valuable tool that can aid health providers and policy makers in understanding public response to health interventions in the ongoing global health crisis. This could include identifying subgroups that are inadequately reached by existing campaigns, as well as predictive modeling of responses to public health messaging to aid health organizations in designing and optimizing outreach campaigns.

## Data Availability

All Tweet IDs associated with this work have been made available via a publicly-accessible repository

https://github.com/TheRensselaerIDEA/COVID-masks-nlp

## Acknowledgements

This study was supported by the Rensselaer Institute for Data Exploration and Applications, the Data INCITE Lab, and a grant from the United Health Foundation.

## Footnotes

1 In addition to explicit COVID-19 keywords such as “coronavirus”, we include keywords such as “school” and “cancelled” in order to include tweets about a wider array of topics impacted by the pandemic

2 Policy can be found at https://developer.twitter.com/en/developer-terms/agreement-and-policy

3 Available at https://therensselaeridea.github.io/COVID-masks-nlp/paper_supplement.pdf

